# Exposure to Anti-Tobacco Messages and Its Association with Tobacco Use Among Zambian Adolescents: Analysis of the 2021 Global Youth Tobacco Survey

**DOI:** 10.64898/2026.02.03.26345513

**Authors:** Davie Edings Mokhola, Wingston Felix Ng’ambi, Chigere Adoration, Cosmas Zyambo

**Affiliations:** Data Science and Statistical Consulting Centre, Lilongwe, Malawi; Health Economics Policy Unit, Department of health Systems and Policy, Kamuzu University of Health Sciences, Lilongwe, Malawi; Department of Public Health and Family Medicine, University of Zambia, Lusaka, Zambia

**Keywords:** Zambia, adolescents, anti-tobacco messages, exposure, mass media, school-based education, tobacco control, WHO FCTC

## Abstract

**Introduction:** Adolescence is a vulnerable period for tobacco initiation, yet evidence on exposure to anti-tobacco messaging in Zambia is limited. Understanding who is reached by media and school-based campaigns is fundamental to designing effective strategies for preventing tobacco initiation and reducing use, in line with tobacco control goals.

**Methods:** We analyzed data from 6,499 in-school adolescents (11–17 years) in the 2021 Zambia Global Youth Tobacco Survey. Exposure was defined as having seen or heard anti-tobacco messages via television, radio, newspapers, or magazines. Weighted prevalence estimates were calculated, and multivariable logistic regression examined associations with sociodemographic, behavioral, and household factors.

**Results:** Overall, 61.6% of adolescents reported exposure to anti-tobacco messages, predominantly through television and radio rather than print media. Exposure was greater among adolescents who had ever tried smoking (OR = 1.37, 95% CI: 1.15–1.63), had friends who smoke (OR = 1.50, 95% CI: 1.27–1.76), lived with a parent/guardian who smokes (OR = 1.58, 95% CI: 1.31–1.93), or received school-based tobacco education (OR = 1.81, 95% CI: 1.60–2.04). Male adolescents had lower odds of exposure (OR = 0.83, 95% CI: 0.73–0.94). Age, grade, current smoking, and observing teachers smoking were not significant predictors.

**Conclusion:** Anti-tobacco messages reached almost two-thirds of Zambian adolescents through schools and mass media, but exposure is strongly influenced by social and household environments. Gender- and context-sensitive strategies targeting male adolescents and peer/family networks could strengthen preventive impact, supporting Zambia’s tobacco control goals and compliance with the WHO Framework Convention on Tobacco Control.

## INTRODUCTION

Tobacco use remains one of the leading causes of preventable disease and premature mortality worldwide. Globally, an estimated 8.7 million deaths each year are attributable to tobacco use, and nearly 90% of smokers initiate the habit during adolescence^1^. Adolescents are particularly vulnerable due to peer influence, risk-taking behaviors, and aggressive marketing strategies that position tobacco as socially desirable^2,3^. Sub-Saharan Africa, once considered a region with low smoking prevalence, is now a major focus for tobacco industry expansion, with projections showing a sharp increase in youth tobacco uptake by 2030 if targeted interventions are not strengthened ^4^.

Zambia is among Africa’s top tobacco-producing countries and is simultaneously experiencing growing youth tobacco experimentation. Findings from the Global Youth Tobacco Survey (GYTS) indicate that 23.2% of Zambian adolescent students have ever smoked cigarettes or other type of tobacco, and 12.9% are current users, raising concerns for future addiction and long-term health consequences^5^. Despite Zambia’s commitment to the WHO Framework Convention on Tobacco Control (WHO FCTC), exposure to tobacco advertising and social acceptability of smoking remain challenges.^6,7^

A key strategy mandated under Article 12 and Article 13 of the FCTC is mass media health communication^8–10^. Anti-tobacco messages delivered through television, radio, print media, and school-based education aim to increase awareness of the harms of tobacco and discourage youth uptake. Evidence from multiple settings suggests that adolescents exposed to anti-tobacco communication are less likely to initiate smoking and more likely to hold negative attitudes toward tobacco use^11,12^. However, exposure may be uneven across demographic groups and may be higher among youth already involved with tobacco or those in social environments supportive of smoking.

While previous analyses of GYTS data in Zambia have described smoking prevalence, limited evidence exists on whether anti-tobacco messages are reaching the intended target population, and whether their exposure is associated with tobacco-related behaviors and social influences. Understanding which groups are exposed and which are not can guide more equitable and effective communication strategies tailored to Zambia’s cultural and policy environment.

Therefore, this study investigates exposure to anti-tobacco messages among in-school adolescents in Zambia using the 2021 Global Youth Tobacco Survey data. Specifically, the study estimates the prevalence of exposure to anti-tobacco messages across different age groups, grades, regions, and behavioral risk categories. It also assesses the association between exposure to anti-tobacco messages and tobacco-related behavioral and environmental factors, while adjusting for sociodemographic variables. By identifying sociodemographic and behavioral characteristics linked to exposure, this study provides insights into opportunities for strengthening youth-focused health communication under Zambia’s tobacco control framework. Findings will support Ministry of Health efforts toward achieving national health goals and protecting future generations from tobacco-related harm.

## METHODS

### Study Design and Data Source

This study utilized data from the 2021 Zambia Global Youth Tobacco Survey (GYTS), a cross-sectional, school-based survey designed to generate nationally representative estimates of tobacco-related indicators among students aged 11–17 years. The GYTS employs a standardized methodology developed by the World Health Organization (WHO) and the Centers for Disease Control and Prevention (CDC)^13,14^. A two-stage cluster sampling design was used, with schools selected proportional to enrollment size in the first stage and classes randomly selected within chosen schools in the second stage. All students in sampled classes were eligible to participate.

### Study Population

A total of 6,499 in-school adolescents with complete information on anti-tobacco message exposure were included in this analysis. Survey weights, primary sampling units (PSUs), and stratification variables were applied to account for the complex sampling design and non-response^13,14^.

### Data Management

Data management involved defining the outcome variable, coding covariates, and handling missing data to ensure consistency and accuracy for analysis^15^. The primary outcome of interest, exposure to anti-tobacco messages, was constructed as a binary variable. Adolescents were coded as exposed (1) if they reported having seen or heard anti-tobacco messages on television or radio (CR30 = 1) and/or in newspapers or magazines (CR31 = 1). Those who did not report exposure on either medium were coded as not exposed (0). Socio-demographic covariates were defined a priori and included in all adjusted models. Age (CR1) was categorized into seven groups: 11 years or younger, 12, 13, 14, 15, 16, and 17 years or older. Sex (CR2) was coded as male or female. Grade level (ZAR3) was classified as Grade 7, 8, or 9. Region was derived from the survey stratum variable and categorized into Lusaka, Tobacco Regions, and the Rest of the Country (ROC)^5^.

Behavioral and environmental predictors were recoded as binary variables (Yes/No) and included: ever tried cigarette smoking (CR5), current smoking (from CR7, CR14, CR43), parental or guardian smoking in the home (OR15), having friends who smoke (ZAR55), observing teachers smoking at school (OR60), and receiving school-based instruction on the dangers of tobacco use (CR33). Missing data were managed by excluding cases missing on the outcome variable. Predictors with missing values were handled using listwise deletion during regression modeling. All variables were checked for consistency and recoded where necessary to ensure alignment with survey documentation prior to analysis.

### Statistical Analysis

Data analyses were performed using R 4.5.2 (R Foundation for Statistical Computing, Vienna, Austria). Data were analyzed using survey-weighted procedures to incorporate clustering and stratification effects. Weighted prevalence estimates with 95% confidence intervals (CIs) were calculated for exposure to anti-tobacco messages overall and across sub-groups. Associations between exposure and predictor variables were investigated using logistic regression^16^. Both Crude models, where each predictor was separately analyzed, and Adjusted model, which included all predictors were used. Age, sex, grade, and region were retained a priori. Stepwise selection based on Akaike Information Criterion (AIC) was applied for optimized model fit. Statistical significance was determined at p < 0.05.

### Ethical Considerations

This study was conducted in accordance with internationally accepted ethical standards for research involving human participants. Access to the anonymized 2021 Global Youth Tobacco Survey (GYTS) dataset was obtained through the World Health Organization (WHO) NCD Microdata Repository. The GYTS data are fully de-identified prior to release, ensuring that no individual respondents can be identified. As the analysis involved secondary use of existing anonymized data and required no direct contact with participants, additional ethical approval or informed consent was not required. The study therefore posed minimal risk and complied with WHO data access and research ethics requirements.

## RESULTS

### Adolescent characteristics

Table 1 shows the demographic, behavioral, and environmental characteristics of the 6,499 in-school adolescents included in the analysis. Adolescents aged 13 to 15 years accounted for the largest proportions of the sample, with 17% aged 13 years, 25% aged 14 years, and 21% aged 15 years. Those aged 11 years or younger represented 4.1% of the sample, while 11% were 17 years or older. Slightly more than half of the participants were female (57%) compared to males (43%). Students were relatively evenly distributed across grade levels, with 35% in Grade 7, 32% in Grade 8, and 33% in Grade 9. Overall, 4,022 students (62%) reported exposure to anti-tobacco messages through television, radio, or newspapers/magazines. Regarding tobacco-related behaviors and environmental factors, 21% of adolescents had ever tried cigarette smoking, 27% were current smokers, 25% had friends who smoke tobacco, and 16% reported that at least one parent or guardian smoked in the home. With respect to school-related factors, 47% of students had been taught about the dangers of tobacco use, and 14% had observed teachers smoking on school premises.

**Table 1:** Characteristics of the adolescents that were assessed on their exposure to anti-tobacco messages in Zambia in 2021.

| Characteristic | N = 6,499 |
| --- | --- |
| **Age, years** |  |
| 11 or younger | 265 (4.1%) |
| 12 | 457 (7.1%) |
| 13 | 1,076 (17%) |
| 14 | 1,589 (25%) |
| 15 | 1,363 (21%) |
| 16 | 1,027 (16%) |
| 17 or older | 678 (11%) |
| **Sex** |  |
| Female | 3,681 (57%) |
| Male | 2,725 (43%) |
| **Grade** |  |
| Grade 7 | 2,271 (35%) |
| Grade 8 | 2,051 (32%) |
| Grade 9 | 2,081 (33%) |
| Exposure to anti-tobacco messages | 4,022 (62%) |
| Ever tried cigarette smoking | 1,083 (21%) |
| Parent/guardian smokes in home | 1,029 (16%) |
| Taught about dangers of tobacco use | 3,004 (47%) |
| Saw teachers smoking at school | 887 (14%) |
| Friends smoke tobacco | 1,609 (25%) |
| Current smoking | 1,723 (27%) |

### Exposure of adolescents to anti-tobacco messages

Table 2 shows the weighted prevalence of exposure to anti-tobacco messages among Zambian adolescents by key demographic, behavioral, and environmental variables. Overall, the prevalence of exposure was 61.62% (95% CI: 58.68–64.56%). By age, prevalence ranged from 60.67% among 14-year-olds to 63.02% among 12-year-olds. Adolescents who were current smokers had a prevalence of 63.15%, compared to 61.43% among non-current smokers. Those who had ever tried cigarette smoking reported a higher prevalence of exposure (68.88%) than those who had never tried smoking (61.19%). Exposure was also higher among adolescents with friends who smoke (68.49%) compared to those without friends who smoke (59.80%), and among those living with a parent or guardian who smokes (70.46%) versus those without (60.48%). By grade level, prevalence ranged from 61.38% in Grade 7 to 63.29% in Grade 8. Slight differences were observed by sex, with 62.81% of females and 61.03% of males reporting exposure. Adolescents who had been taught about the dangers of tobacco use in school reported higher exposure (69.21%) than those who had not received such instruction (55.72%). Exposure among adolescents who had seen teachers smoking at school was 64.26%, compared to 61.70% among those who had not. Across the surveyed region (ROC), the prevalence of exposure was 61.89%. All prevalence estimates were weighted to account for the complex survey design, including stratification and clustering.

**Table 2:** Prevalence of exposure to anti-tobacco messages amongst the adolescents in Zambia in 2021.

| **Characteristic** | **Weighted % (95% CI)** |
| --- | --- |
| **Age, years** |  |
| 11 or younger | 61.13 (55.25 - 67.01) |
| 12 | 63.02 (58.59 - 67.45) |
| 13 | 61.90 (58.99 - 64.80) |
| 14 | 60.67 (58.26 - 63.07) |
| 15 | 62.14 (59.57 - 64.72) |
| 16 | 62.90 (59.95 - 65.86) |
| 17 or older | 62.68 (59.04 - 66.33) |
| **Current smoking** |  |
| No | 61.43 (60.05 - 62.81) |
| Yes | 63.15 (60.87 - 65.42) |
| **Ever tried cigarette smoking** |  |
| No | 61.19 (59.68 - 62.71) |
| Yes | 68.88 (66.12 - 71.64) |
| **Friends smoke tobacco** |  |
| No | 59.80 (58.42 - 61.18) |
| Yes | 68.49 (66.22 - 70.76) |
| **Grade** |  |
| Grade 7 | 61.38 (59.38 - 63.39) |
| Grade 8 | 63.29 (61.20 - 65.37) |
| Grade 9 | 61.60 (59.51 - 63.70) |
| **Parent/guardian smokes in home** |  |
| No | 60.48 (59.18 - 61.78) |
| Yes | 70.46 (67.67 - 73.25) |
| **Saw teachers smoking at school** |  |
| No | 61.70 (60.42 - 62.97) |
| Yes | 64.26 (61.11 - 67.42) |
| **Sex** |  |
| Female | 62.81 (61.25 - 64.37) |
| Male | 61.03 (59.20 - 62.86) |
| **Taught about dangers of tobacco use** |  |
| No | 55.72 (54.05 - 57.38) |
| Yes | 69.21 (67.56 - 70.86) |

### Media for anti-tobacco messages

**Figure 1.** illustrates the distribution of exposure to anti-tobacco messages among Zambian adolescents by media type and selected demographic characteristics. For television and radio (CR30), exposure was relatively uniform across age bands, ranging from 44.1% among adolescents aged 11–12 years to 44.8% among those aged 13–14 years, with 44.5% among those aged 15–17 years (Panel A). No substantial differences were observed by sex, with 46.0% of both females and males reporting exposure (Panel B). By grade, exposure ranged from 46.0% in Grade 7 to 45.3% in Grade 9 (Panel C).

**Figure 1.**
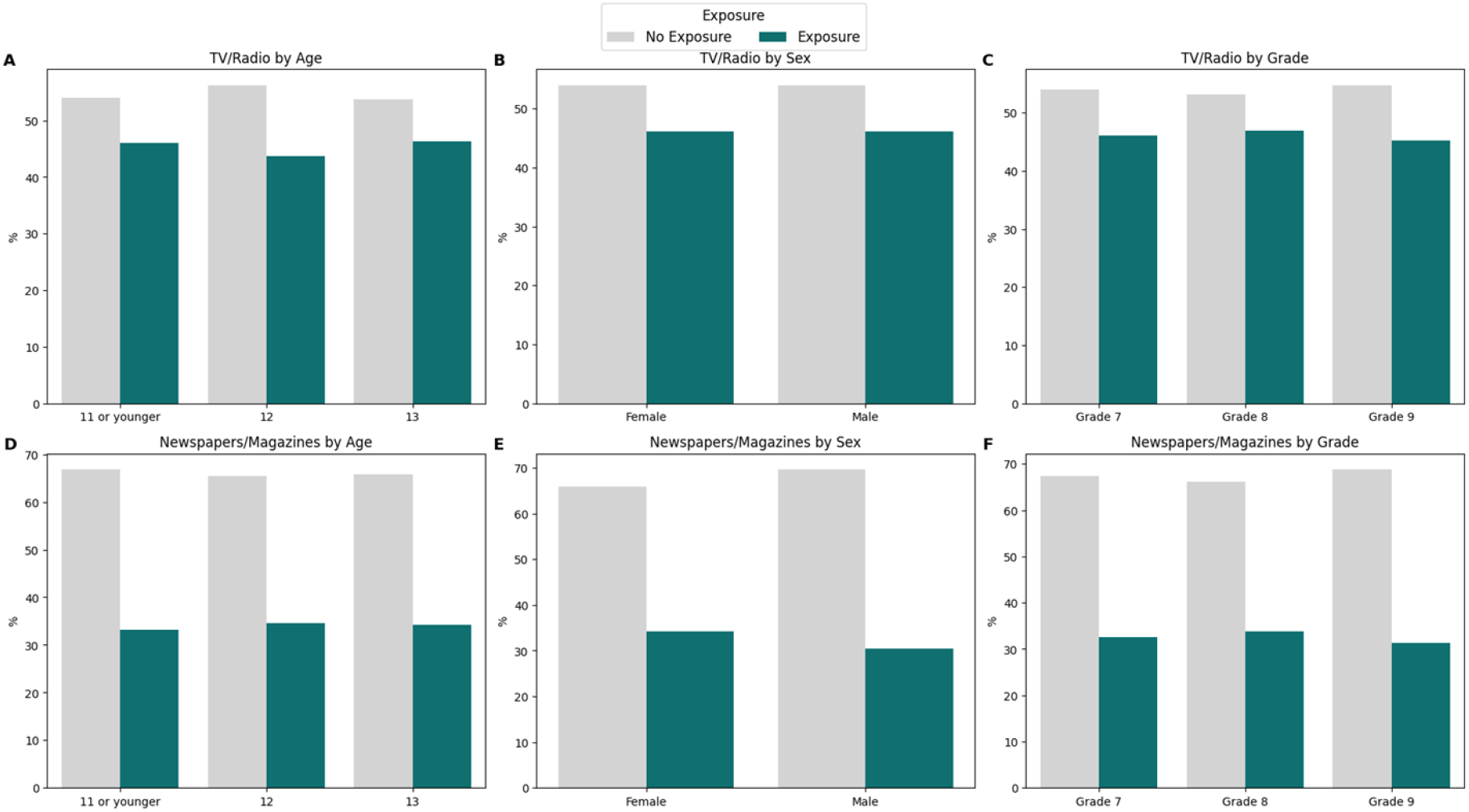
Exposure to Anti-Tobacco Messages Among Zambian Adolescents by Media Type and Selected Demographic Characteristics in 2021. Figure 1. Weighted proportions of Zambian adolescents exposed to anti-tobacco messages by media type and demographic characteristics. Panels A–C show TV/radio (CR30) exposure by age, sex, and grade. Panels D–F show newspapers/magazines (CR31) exposure by age, sex, and grade. Percentages account for survey weights, stratification, and clustering.

Exposure via newspapers and magazines (CR31) was lower overall. Age-specific exposure ranged from 30.9% among adolescents aged 15–17 years to 34.2% among those aged 13–14 years, with 31.8% among 11–12-year-olds (Panel D). Small differences by sex were observed, with 34.2% of females and 30.4% of males reporting exposure (Panel E). Across grades, exposure ranged from 32.6% in Grade 7 to 31.2% in Grade 9 (Panel F). These findings indicate that television and radio were the predominant channels reaching adolescents, while print media reached a smaller proportion. Patterns of exposure were generally similar across age bands and grade levels, with minor sex differences noted for print media.

### Factors Associated with Anti-Tobacco Message Exposure

Figure 2 shows regression results on the factors associated with exposure to anti-tobacco messages among Zambian adolescents. Adolescents who had ever tried cigarette smoking had higher odds of exposure to anti-tobacco messages (OR = 1.37, 95% CI: 1.15–1.63). Similarly, adolescents with friends who smoke had higher odds of exposure (OR = 1.50, 95% CI: 1.27–1.76), as did those living with a parent or guardian who smokes (OR = 1.58, 95% CI: 1.31–1.93). Being taught about the dangers of tobacco use at school was associated with the highest odds of exposure (OR = 1.81, 95% CI: 1.60–2.04). Male adolescents had lower odds of exposure compared to females (OR = 0.83, 95% CI: 0.73–0.94).

**Figure 2:**
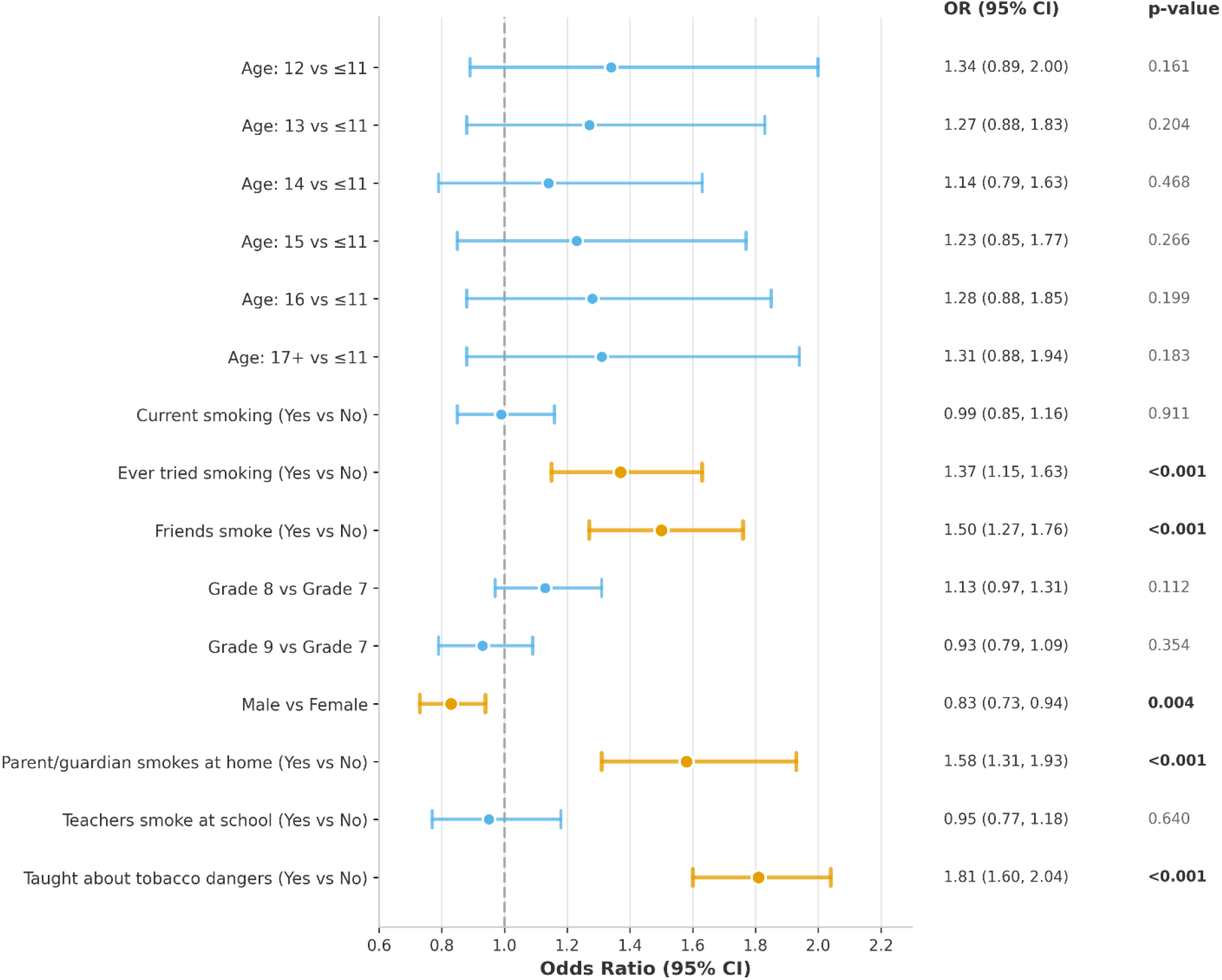
Adjusted Odds Ratios for Exposure to anti-tobacco messages amongst the adolescents in Zambia in 2021

## DISCUSSION

This study examined the prevalence and determinants of exposure to anti-tobacco messages among in-school adolescents in Zambia, providing important insights into how tobacco control communication reaches young people within a high-risk developmental period. Overall, nearly two-thirds of adolescents reported exposure to anti-tobacco messages through mass media or print sources, indicating moderate penetration of tobacco control messaging. This level of exposure is comparable to findings from other low- and middle-income countries (LMICs), where school-based education and mass media campaigns constitute the backbone of adolescent tobacco prevention efforts ^10,17^.

The descriptive findings show that a substantial proportion of adolescents were already exposed to tobacco-related risk environments, including peer smoking, parental smoking, and prior experimentation with cigarettes. Similar patterns have been documented globally and across sub-Saharan Africa, where early exposure to smoking within social networks increases both susceptibility to tobacco use and awareness of tobacco-related messaging^17^. The relatively high overall exposure observed in this study suggests that anti-tobacco messages may be reaching adolescents who are most vulnerable to tobacco initiation.

Prevalence estimates further demonstrated that exposure to anti-tobacco messages was consistently higher among adolescents who had ever tried smoking, those with friends who smoke, and those living with a parent or guardian who smokes. This pattern aligns with evidence from global Youth Tobacco Surveys showing that adolescents embedded in smoking environments are more likely to recall or notice anti-tobacco messaging, possibly due to increased salience or targeted interventions^18^. However, this also raises concerns that adolescents without direct exposure to smoking risks may be less effectively reached, potentially limiting the preventive impact of current communication strategies.

Exposure to anti-tobacco messages among Zambian adolescents was higher through television and radio compared to print media, suggesting that broadcast channels remain the most effective means of reaching this population^19^. Exposure was generally consistent across age bands and grades, with slightly higher print media exposure among females. These patterns are consistent with findings from other LMICs, where broadcast campaigns typically achieve wider reach and gender differences in message reception are observed^20,21^. The results highlight the continued importance of mass media in adolescent tobacco prevention, while suggesting the need to tailor print media to improve engagement across all groups.

School-based education emerged as a central determinant of exposure. Adolescents who reported being taught about the dangers of tobacco use had substantially higher prevalence of exposure and the strongest association in the adjusted regression model. This finding is consistent with extensive global and African literature demonstrating that school-based tobacco education improves awareness and recall of anti-tobacco messages, even in settings where enforcement of broader tobacco control policies may be weak^22,23^. It reinforces the role of schools as a critical platform for tobacco prevention among adolescents.

The regression analysis highlights the importance of social and household contexts. Adolescents with friends who smoke and those living with parents or guardians who smoke had significantly higher odds of exposure to anti-tobacco messages. These findings support socio-ecological models of health behavior, which emphasize the influence of interpersonal and household environments on both health risks and exposure to health information^24–26^. Similar associations have been reported in studies from South Africa and Ghana, where peer and parental smoking increased adolescents’ engagement with tobacco-related messaging^27,28^

Gender differences were also evident, with male adolescents having lower odds of exposure compared to females. This mirrors findings from other African and global studies suggesting that girls may be more engaged with school-based health education or more receptive to health promotion messages^29–31^. Given that tobacco use prevalence is often higher among males, this gap highlights the need for gender-responsive communication strategies.

Age, grade level, current smoking status, and observing teachers smoking at school were not significantly associated with exposure after adjustment. The lack of association with age and grade suggests relatively uniform delivery of anti-tobacco messaging across schooling levels, likely reflecting standardized curricula and mass media approaches. The non-significant association with current smoking aligns with global evidence that exposure to information alone may be insufficient to distinguish smokers from non-smokers, emphasizing the need for comprehensive tobacco control measures beyond awareness campaigns^32,33^

From a policy perspective, these findings are highly relevant to Zambia’s commitments under the WHO Framework Convention on Tobacco Control, particularly Articles 12 and 13 on education, public awareness, and communication. The strong role of school-based education aligns with Zambia’s national adolescent health and non-communicable disease strategies.^34,35^ However, the influence of peer and household smoking highlights the need to complement school and media interventions with family- and community-based approaches to strengthen overall impact.

## CONCLUSION

This study demonstrates that exposure to anti-tobacco messages among in-school adolescents in Zambia is relatively widespread, with nearly two-thirds reporting exposure through mass media or school-based channels. Exposure was more common among adolescents who had ever tried smoking, those with friends or parents who smoke, and those who had received school-based instruction on the dangers of tobacco use. Female adolescents were more likely to report exposure than males, while age, grade level, current smoking status, and observing teachers smoking at school were not significantly associated with exposure. These findings highlight the important role of schools and mass media in disseminating anti-tobacco messages, while also underscoring the influence of social and household environments on message reach. To strengthen adolescent tobacco prevention in Zambia, future efforts should prioritize more targeted communication strategies that better reach male adolescents and extend beyond schools to address peer and family contexts, in alignment with national tobacco control policies and the WHO Framework Convention on Tobacco Control.

## Data Availability

The data can be accessed from NCD Monitor. We have made available the code used in the analysis.

## REFERENCES

1. Murray, C. J. L. et al. Global burden of 87 risk factors in 204 countries and territories, 1990–2019: a systematic analysis for the Global Burden of Disease Study 2019. The Lancet 396, 1223–1249 (2020).

2. Peers and adolescent smoking - Kobus - 2003 - Addiction - Wiley Online Library. https://onlinelibrary.wiley.com/doi/abs/10.1046/j.1360-0443.98.s1.4.x.

3. Frontiers | Adolescents’ own views on their risk behaviors, and the potential effects of being labeled as risk-takers: A commentary and review. https://www.frontiersin.org/journals/psychology/articles/10.3389/fpsyg.2022.945775/full.

4. (PDF) Tobacco Use in Sub-Saharan Africa: The Risks and Challenges. https://www.researchgate.net/publication/333293410_Tobacco_Use_in_Sub-Saharan_Africa_The_Risks_and_Challenges.

5. Zambia 2021 Global Youth Tobacco Survey (GYTS) Country Report | WHO | Regional Office for Africa. https://www.afro.who.int/countries/zambia/publication/zambia-2021-global-youth-tobacco-survey-gyts-country-report.

6. The WHO Framework Convention on Tobacco Control: 10 Years of Implementation in the African Region. (World Health Organization, Regional Office for Africa, Brazzaville, Republic of Congo, 2015).

7. Labonté, R. et al. The institutional context of tobacco production in Zambia. Glob. Health 14, 5 (2018).

8. FCTC Regulations on the Need to Protect Public Health Policies from Tobacco Industry Interference. Tobacco Tactics https://www.tobaccotactics.org/article/fctc-regulations-protect-public-health-policies-interference/.

9. Nagler, R. H. & Viswanath, K. Implementation and Research Priorities for FCTC Articles 13 and 16: Tobacco Advertising, Promotion, and Sponsorship and Sales to and by Minors. Nicotine Tob. Res. 15, 832–846 (2013).

10. Indicator Metadata Registry Details. https://www.who.int/data/gho/indicator-metadata-registry/imr-details/2266.

11. Addo, I. Y. et al. Exposure to pro-tobacco and anti-tobacco media messages and events and smoking behaviour among adolescents in Gambia. BMC Public Health 24, 1041 (2024).

12. Adolescents’ Perceptions of Tobacco Control Measures in the United Kingdom - Abraham Brown, Crawford Moodie, 2012. https://journals.sagepub.com/doi/10.1177/1524839910369222.

13. World Health Organization. Global Youth Tobacco Survey (GYTS). WHO Noncommunicable Disease Surveillance, Monitoring and Reporting https://www.who.int/teams/noncommunicable-diseases/surveillance/systems-tools/global-youth-tobacco-survey (2025).

14. Warren, C. The Global Youth Tobacco Survey (GYTS): Linking data to the implementation of the WHO Framework Convention on Tobacco Control. BMC Public Health 8 Suppl 1, S1 (2008).

15. Ng’ambi, W. F. & Muula, A. S. A reproducible R workflow to preserve variable and value labels in Stata, SPSS, and SAS datasets for transparent and reproducible health research. Malawi Med. J. J. Med. Assoc. Malawi 37, 193–206 (2025).

16. Ng’ambi WF. A Practical Guide to Key Considerations in Logistic Regression. Malawi Med J [Internet]. 2023 [Cited 2025 Oct 27];35(2):135–7. Available from: https://Www.Mmj.Mw/?P=13527.

17. Addo, I. et al. Exposure to pro-tobacco and anti-tobacco media messages and events and smoking behaviour among adolescents in Gambia. BMC Public Health 24, 1–13 (2024).

18. Stephano, E., Rwegoshola, K., Mtoro, M. & Nyundo, A. Using the theory of triadic influence to establish the trend in tobacco use and its correlates among in-school adolescents in Tanzania (2003–2016): findings from the global youth tobacco survey. Discov. Public Health 22, (2025).

19. Sharma, K., Gawde, N. & Pednekar, M. S. Are Anti-Tobacco Messages Delivered through Different Mass-Media Channels Effective in India? Results from GATS-II Survey. Asian Pac. J. Cancer Prev. APJCP 25, 2751–2760 (2024).

20. Hossain, M. M., Bin, S. A. & Kabir, M. A. Determinants of mass media exposure and involvement in information and communication technology skills among Bangladeshi women. Jahangirnagar Univ J Stat Stud 36, 161–172 (2022).

21. Neha Singh, N. S. & TJPRC. A Study on Knowledge of Rural Women Towards Mass Media and Its Usage in Bikaner District. Int. J. Educ. Sci. Res. 8, 63–70 (2018).

22. (PDF) Awareness of tobacco control policies and anti-tobacco attitudes and behaviors among school personnel. https://www.researchgate.net/publication/361201628_Awareness_of_tobacco_control_policies_and_anti-tobacco_attitudes_and_behaviors_among_school_personnel.

23. Chen, N., Dai, L., Wang, J., Zhang, L. & Zhu, J. Changes of campus tobacco control environment and the impact on tobacco control behaviors among secondary school personnel in Shanghai, China. Tob. Induc. Dis. 22, 10.18332/tid/191763 (2024).

24. Lin, M. et al. Factors influencing adolescent experimental and current smoking behaviors based on social cognitive theory: A cross-sectional study in Xiamen. Front. Public Health 11, 1093264 (2023).

25. Ismail, R. et al. Prevalence, risk factors, and protective factors of tobacco use among school-going adolescents at drug-abuse hot-spots in Malaysia. Addict. Behav. 171, 108455 (2025).

26. Influence of smoking by family and best friend on adolescent tobacco smoking: results from the 2002 New Zealand national survey of Year 10 students | Request PDF. https://www.researchgate.net/publication/6160573_Influence_of_smoking_by_family_and_best_friend_on_adolescent_tobacco_smoking_results_from_the_2002_New_Zealand_national_survey_of_Year_10_students.

27. Logo, D. D. et al. Profile and predictors of adolescent tobacco use in Ghana: evidence from the 2017 Global Youth Tobacco Survey (GYTS). J. Prev. Med. Hyg. 62, E664–E672 (2021).

28. Morrison, R. A. Parental, Peer, and Tobacco Marketing Influences on Adolescent Smoking in South Africa.

29. Magadi, M., Kaseje, D., Wafula, C., et al. (2021). Sexual and reproductive health knowledge and behaviour of adolescent boys and girls aged 10–19 years in western Kenya. Journal of Biosocial Science. - Google Search. https://www.google.com/search?q=Magadi%2C+M.%2C+Kaseje%2C+D.%2C+Wafula%2C+C.%2C+et+al.+(2021).+Sexual+and+reproductive+health+knowledge+and+behaviour+of+adolescent+boys+and+girls+aged+10%E2%80%9319+years+in+western+Kenya.+Journal+of+Biosocial+Science.&rlz=1C1AJCO_enNG1194NG1194&oq=Magadi%2C+M.%2C+Kaseje%2C+D.%2C+Wafula%2C+C.%2C+et+al.+(2021).+Sexual+and+reproductive+health+knowledge+and+behaviour+of+adolescent+boys+and+girls+aged+10%E2%80%9319+years+in+western+Kenya.+Journal+of+Biosocial+Science.&gs_lcrp=EgZjaHJvbWUyBggAEEUYOdIBBzg5OGowajeoAgCwAgA&sourceid=chrome&ie=UTF-8.

30. Achak, D., Azizi, A., et al. (2024). The health behaviors differences among male and female school-age adolescents in the Middle East and North Africa region countries: A meta-analysis of the GSHS data. Frontiers in Public Health. Frontiers - Google Search. https://www.google.com/search?q=Achak%2C+D.%2C+Azizi%2C+A.%2C+et+al.+(2024).+The+health+behaviors+differences+among+male+and+female+school-age+adolescents+in+the+Middle+East+and+North+Africa+region+countries%3A+A+meta-analysis+of+the+GSHS+data.+Frontiers+in+Public+Health.+Frontiers&rlz=1C1AJCO_enNG1194NG1194&oq=Achak%2C+D.%2C+Azizi%2C+A.%2C+et+al.+(2024).+The+health+behaviors+differences+among+male+and+female+school-age+adolescents+in+the+Middle+East+and+North+Africa+region+countries%3A+A+meta-analysis+of+the+GSHS+data.+Frontiers+in+Public+Health.+Frontiers&gs_lcrp=EgZjaHJvbWUyBggAEEUYOdIBBzc1NmowajmoAgawAgHxBZfyR3qXm837&sourceid=chrome&ie=UTF-8.

31. Tian, H., Dang, R., & Chen, J. (2024). Associations between health promotion behaviors and multidimensional health: Gender differences among middle-school students. Journal of Men’s Health. - Google Search. https://www.google.com/search?q=Tian%2C+H.%2C+Dang%2C+R.%2C+%26+Chen%2C+J.+(2024).+Associations+between+health+promotion+behaviors+and+multidimensional+health%3A+Gender+differences+among+middle-school+students.+Journal+of+Men%E2%80%99s+Health.&rlz=1C1AJCO_enNG1194NG1194&oq=Tian%2C+H.%2C+Dang%2C+R.%2C+%26+Chen%2C+J.+(2024).+Associations+between+health+promotion+behaviors+and+multidimensional+health%3A+Gender+differences+among+middle-school+students.+Journal+of+Men%E2%80%99s+Health.&gs_lcrp=EgZjaHJvbWUyBggAEEUYOdIBBzc3NmowajmoAgawAgHxBQSX7KqaL0wu&sourceid=chrome&ie=UTF-8xwid.

32. U.S. National Library of Medicine. Multilevel analysis of school anti-smoking education and current cigarette use among South African students. PubMed. (2017) -Google Search. https://www.google.com/search?q=U.S.+National+Library+of+Medicine.+Multilevel+analysis+of+school+anti-smoking+education+and+current+cigarette+use+among+South+African+students.+PubMed.+(2017)&rlz=1C1AJCO_enNG1194NG1194&oq=U.S.+National+Library+of+Medicine.+Multilevel+analysis+of+school+anti-smoking+education+and+current+cigarette+use+among+South+African+students.+PubMed.+(2017)&gs_lcrp=EgZjaHJvbWUyBggAEEUYOdIBBzcwNWowajmoAgawAgHxBXthrNWpDTkA&sourceid=chrome&ie=UTF-8.

33. Wiehe, S., Garrison, M., Christakis, D., Ebel, B. & Rivara, F. A systematic review of school-based prevention trials with long-term follow-up. J. Adolesc. Health Off. Publ. Soc. Adolesc. Med. 36, 162–9 (2005).

34. Government of the Republic of Zambia. National Adolescent Health Strategic Plan 2022–2026. Ministry of Health. (2022) -Google Search. https://www.google.com/search?q=Government+of+the+Republic+of+Zambia.+National+Adolescent+Health+Strategic+Plan+2022%E2%80%932026.+Ministry+of+Health.+(2022)&rlz=1C1AJCO_enNG1194NG1194&oq=Government+of+the+Republic+of+Zambia.+National+Adolescent+Health+Strategic+Plan+2022%E2%80%932026.+Ministry+of+Health.+(2022)&gs_lcrp=EgZjaHJvbWUyBggAEEUYOdIBBzc1NWowajSoAgCwAgE&sourceid=chrome&ie=UTF-8.

35. Zambia national health strategic plan 2022–2026: Towards attainment of quality universal health coverage through decentralization | GPC. https://hivpreventioncoalition.unaids.org/en/resources/zambia-national-health-strategic-plan-2022-2026-towards-attainment-quality-universal.

